# Clinical evaluation of IFN beta1b in COVID-19 pneumonia: a retrospective study

**DOI:** 10.1101/2020.05.15.20084293

**Authors:** Miriam Estébanez, German Ramírez-Olivencia, Tatiana Mata, David Martí, Carlos Gutierrez, Begoña de Dios, María Dolores Herrero, Ana Roel, Yolanda Martínez, Alejandro Aguirre, Francisco Alcántara-Nicolás, Pablo Fernández-González, Elena López, Lucía Elena Ballester, María Mateo-Maestre, Sergio Campos, María Jesús Sánchez-Carrillo, Antonio Fe, Francisco Javier Membrillo de Novales, COVID 19 CENTRAL DEFENSE HOSPITAL “GÓMEZ ULLA” TEAM

**Affiliations:** Central Defense Hospital “Gómez Ulla”, Glorieta Ejército, 1, 28047 Madrid (Spain)

## Abstract

**Background:** COVID-19 pneumonia is associated with significant mortality and has no approved antiviral therapy. Interferon beta1 has shown in vitro studies a potent inhibition of SARS-CoV and MERS-CoV. In an *in vitro* study, SARS-CoV-2 had more sensitivity to IFN-I pretreatment that SARS-CoV. A combination of IFN beta1b administered subcutaneously with other antiviral treatments has been recommended in several guidelines. However, clinical trial results for the treatment of COVID-19 are pending. We aimed to assess the efficiency of IFN beta1b in COVID19 comparing the in-hospital mortality between patients who received IFN beta1b and patients did not receive.

**Methods:** In this retrospective cohort study, we included hospitalized adults with COVID-19 between February 23th and April 4th, 2020, at the Central Defense Hospital (Madrid, Spain). Subcutaneous interferon beta-1b was recommended in moderate-severe pneumonia. The primary endpoint was in-hospital mortality. Univariate and multivariate analysis was performed to identify variables associated with in-hospital mortality.

**Findings:** We analyzed 256 patients (106 patients in interferon group and 150 patients in control group). At admission, patients who did not receive interferon beta1b presented a greater number of comorbidities. The overall mortality rate was 24.6% (63/256). Twenty-two patients (20.8%) in the interferon group died and 41 (27.3%) in the control group (p=0.229). In the multivariate analysis, the predictors of in-hospital mortality were age, severity of clinical picture at admission and hydroxychloroquine treatment.

**Interpretation:** In hospitalized patients with COVID-19, interferon beta1b treatment was not associated to decrease in-hospital mortality. Further assessment of the earlier administration of this drug in randomized trials is recommended.

**Funding:** none.

**RESEARCH IN CONTEXT:** *Evidence before this study:* We searched Pubmed on April 27^th^, 2020, for articles evaluating the efficacy of interferon beta in patients infected with severe acute respiratory syndrome coronavirus 2 (SARS-CoV-2), using the terms: “interferon beta and (COVID-19 or SARS-CoV-2)”. We only found 5 articles. Of them, there was only one original article in English, which was a descriptive study of a case series with solid organ transplant from Spain.

*Added value of this study:* This is the first article that reports the efficacy of interferon beta1b in the treatment of patients with COVID-19. We compared the in-hospital mortality between patients who received interferon beta1b and patients who did not. Patients in both groups received other drugs with a potential antiviral and immunomodulatory effect. There was no significant difference in in-hospital mortality between both groups.

*Implications of all the available evidence:* In our retrospective cohort, treatment with interferon beta1b had not impact on in-hospital survival, however it would be of clinical interest to evaluate the effect of early administration of this drug in the control of SARS-CoV-2 infection in larger randomized clinical trials.

## INTRODUCTION

A new respiratory disease was reported to the WHO China Country Office on 31 December 2019 (1). A cluster of cases of pneumonia of unknown etiology had been detected in Wuhan City, Hubei Province of China, causing severe respiratory illness similar to severe acute respiratory syndrome coronavirus and was associated with ICU admission and high mortality in some cases (2). A new type of coronavirus (named lately SARS-CoV-2), was isolated on 7 January 2020 (3). On January 30th, 2020, WHO declared the outbreak of nCoV-2019 as a Public Health Emergency of International Concern (PHEIC) (4). From the initial description, the disease has spread around the world becoming a pandemic threat, and has affected not only health, but economic and social affairs. By April 27th, 2020, there were more than 2.5M total confirmed cases and more than180000 deaths, in 213 countries (5).

Initial descriptions reported flu-like symptoms such as fever, cough, dysnea, headache, myalgias and asthenia. Digestive complains were also reported including nausea, vomiting and diarrhea. With spreading of the disease, there here have been described new clinical features of the disease, such as neurological, myocardial, ophtalmic, cutaneous or thrombotic events. When coronavirus invades the host, a complex intracellular mechanism is activated, and synthesis of type I interferons (IFNs) is promoted. SARS-CoV-2 penetrates alveolar cell (pneumocytes type II) using the receptor for angiotensin-converting enzyme 2 (ACE-2), a membrane exopeptidase. Activating the downstream JAK STAT signal-pathway, IFNs limit virus spread, and play an immunomodulatory role through macrophage, NK cells and T/B cells. Blocking the production of IFNs allows virus to survive (6). On the other hand, coronavirus infection activates the immune system generating an excessive response that could lead to greater lung injury and worse clinical evolution. However, this hyperactivation is insufficient to control the infection and leads to further tissue damage, being able to develop the Adult Respiratory Distress Syndrome (ARDS); the latter has been described as the main cause of mortality from COVID-19. Excessive activation of the innate immune system also causes damage to the microvascular system and activates the coagulation system and inhibition of fibrinolysis, causing sepsis-induced coagulopathy or disseminated intravascular coagulation, with widespread microcirculatory disorders leading to the development of multi-organ failure. Recommendations about treatment, initially based on antiviral drugs, are now considering immunomodulation as a key stone but not the only one. Several different treatments have been used for COVID-19, including lopinavir/ritonavir, interferon, hydroxicloroquine, tocilizumab, baricitinib, immunoglobulin and corticoids with no or conflicting results. No approved treatment is currently available, but there are therapeutic strategies in SARS and MERS-CoV disease on clinical or preclinical research that would suggest interferon as pillar tool in COVID-19.

Antiviral effect of interferon has been described in other positive single-stranded RNA virus diseases such as HCV. Antiviral activity is based on the activation of RNase that breaks-down the nucleic acid chain of the virus. In addition, it blocks the translation of RNA, prevents encapsidation and viral release (7). There are in vitro tests showing activity against MERS-CoV (8) and studies on animal models had promising results (9), but studies to evaluate clinical efficacy had variable outcomes (10 11 12 13).

The aim of this study is to assess the effectivity and safety of interferon beta1b in the treatment of COVID-19 pneumonia.

## METHODS

### Study design and participants

This is a single-center retrospective cohort study. We enrolled all hospitalized adults (aged ≥16 years) at the Central Defense Hospital “Gómez Ulla” (Madrid, Spain) with COVID-19 and had a definitive outcome (dead or discharge) between February 23th and April 4th. COVID-19 diagnosis was defined according to Spanish Ministry of Health definitions on March 19th, 2020. COVID19 case including confirmed cases by reverse-transcriptase–polymerase chain-reaction (RT-PCR) assay for SARS-CoV-2 in a respiratory tract sample and probable cases (bilateral interstitial pneumonia with clinical picture compatible with a COVID-19 diagnosis with no laboratory tests or non-concluding SARS-CoV-2 test). No exclusion criteria were applied at this stage.

### Data collection

Demographical data, comorbidities, treatments prescribed for COVID-19, clinical severity at admission and analytical parameters at admission were extracted from the electronic medical records by two physicians and entered into a database. In case of differences of interpretation a third investigator checked the medical records and adjudicated any difference.

### Definitions

Severity of clinical picture at admission was defined according to Ministry of Health of Spain medical treatment protocol, March 19th, 2020: mild (SpO2 in rest above 93%), moderate (need for supplemental oxygen with fraction of inspired oxygen less than 50%), severe (who required mechanical ventilation or had a fraction of inspired oxygen of at least 50% or more).

Hypertension was defined as previous arterial hypertension requiring any pharmacological treatment. Diabetes Mellitus was defined as previous hyperglycemia requiring any pharmacological hypoglycemic treatment. Dyslipidemia was defined as previous alteration in lipid profile requiring pharmacological treatment. Cardiomyopathy was defined as any previous diagnosis of cardiac chronic disease or acute cardiac event. Respiratory disease was defined as any previous lower respiratory tract chronic disease requiring chronic pharmacological treatment. Cancer was defined as any previously diagnosed malignancy. Dementia was defined as any mental chronic disease altering cognitive capabilities.

### Procedures

Interferon beta1b (Betaferon®) was given by subcutaneous injection at a dose of 250 μg on alternate days. Patients included in the interferon group had received at least one dose. Local treatment protocol recommended interferon beta1b in the hospitalized patients with moderate-severe pneumonia, with a duration between 3-5 doses. The number of doses was at the discretion of the clinician according to the evolution of the patient and tolerance. Patients in both groups were treated with other specific drugs with potential activity against SARS-CoV-2 and/or COVID-19 immune disorders leading to ARDS. These drugs could include antivirals (lopinavir/ritonavir, and/or hydroxychloroquine or chloroquine), and/or anti-inflammatory drugs (steroids and/or tocilizumab). Antibiotics were scheduled only in the case of suspected bacterial infection.

In addition to regular clinical monitoring, renal function, liver enzymes, and blood count were assessed at baseline and closely throughout the treatment course at the discretion of the clinician. Chest radiographs were also done for all inpatients.

### Outcomes

The primary endpoint was in-hospital mortality in two different regimens: with and without interferon betalb.

### Statistical methods

Simple characteristics were described using absolute and relative frequencies for categorical variables and means ±SD or median (IQR) for continuous variables. Chi-square test or the nonparametric Mann-Whitney U-test were used to compare continuous baseline variables. Comparisons of categorical variables between groups were conducted using the Pearson’s chi-squared test or Fisher’s exact test, as appropriate.

Univariate analysis was performed to identify variables associated with mortality. The variables evaluated were: age, sex, presence of comorbidities (hypertension, dyslipidemia, diabetes, lung disease, heart disease, cancer, dementia), number of comorbidities, clinical severity at admission (mild, moderate, severe), laboratory parameters at admission and use of other treatments. Age and laboratory parameters were categorized in according to the ROC curves of each parameter: age (<65 vs. ≥65 years), lymphocytes (<1000 cls./uL vs. ≥1000 cls./uL), C-reactive protein (<14 mg/dl vs. ≥14 mg/dl), D-dimer (<1000 ng/mL vs. ≥1000 ng/mL), ferritin (<1000 ng./mL vs. ≥1000 ng./mL), lactate dehydrogenase (<400U/L vs. ≥400 U/L). A multivariate logistic regression of variables with significance in univariate analysis of p<0,1 was performed. Statistical analysis was performed using SPSS version 25 statistical package. All tests were 2-sided, p values <0.05 were considered significant.

### Ethics approvals

From beginning of COVID-19 epidemic, eligible hospitalized patients were offered treatment with subcutaneous interferon beta1b after informed verbal consent was obtained from the patients themselves or their closest relative. The research protocol was approved by the Ethics Committee on Clinical Investigation of the Ministry of Defense of Spain (code 25/20).

### Role of funding source

No external funding was received for this study.

## RESULTS

A total of 256 patients with COVID19 were admitted in our hospital, and died or have been discharged between February 23th and April 4th. Most patients were male (59.4%) with mean age of 63.7 years. The most frequent comorbidity was hypertension (44.5%), followed by dyslipemia and cardiopathy (Table 1). Overall patients included, 61.2% presented at least two comorbidities. Median time from illness onset to hospital admission was 7 days (IQR 3-9). At admission, 46.3% of patients had a mild clinical picture, 36.1% moderate and 17.6% severe. At hospital discharge, 89.5% of patients has been diagnosed with pneumonia. Most of the patients in our cohort received hydroxychloroquine (77%), followed by azythromycin (62.9%), interferon beta1b (41.4%), lopinavir/ritonavir (36.1%), and corticosteroids (25.8%).

**Table 1.**
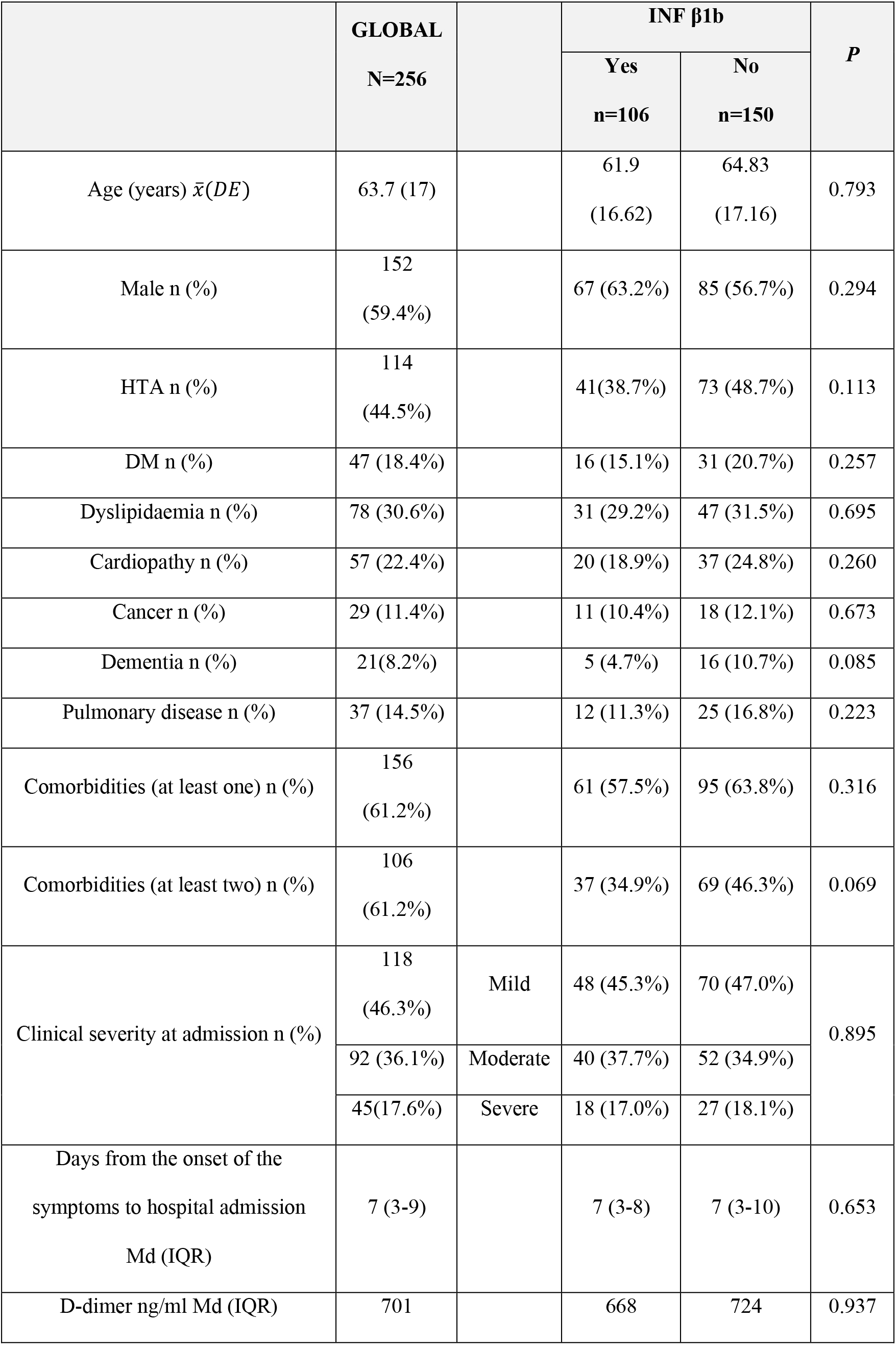
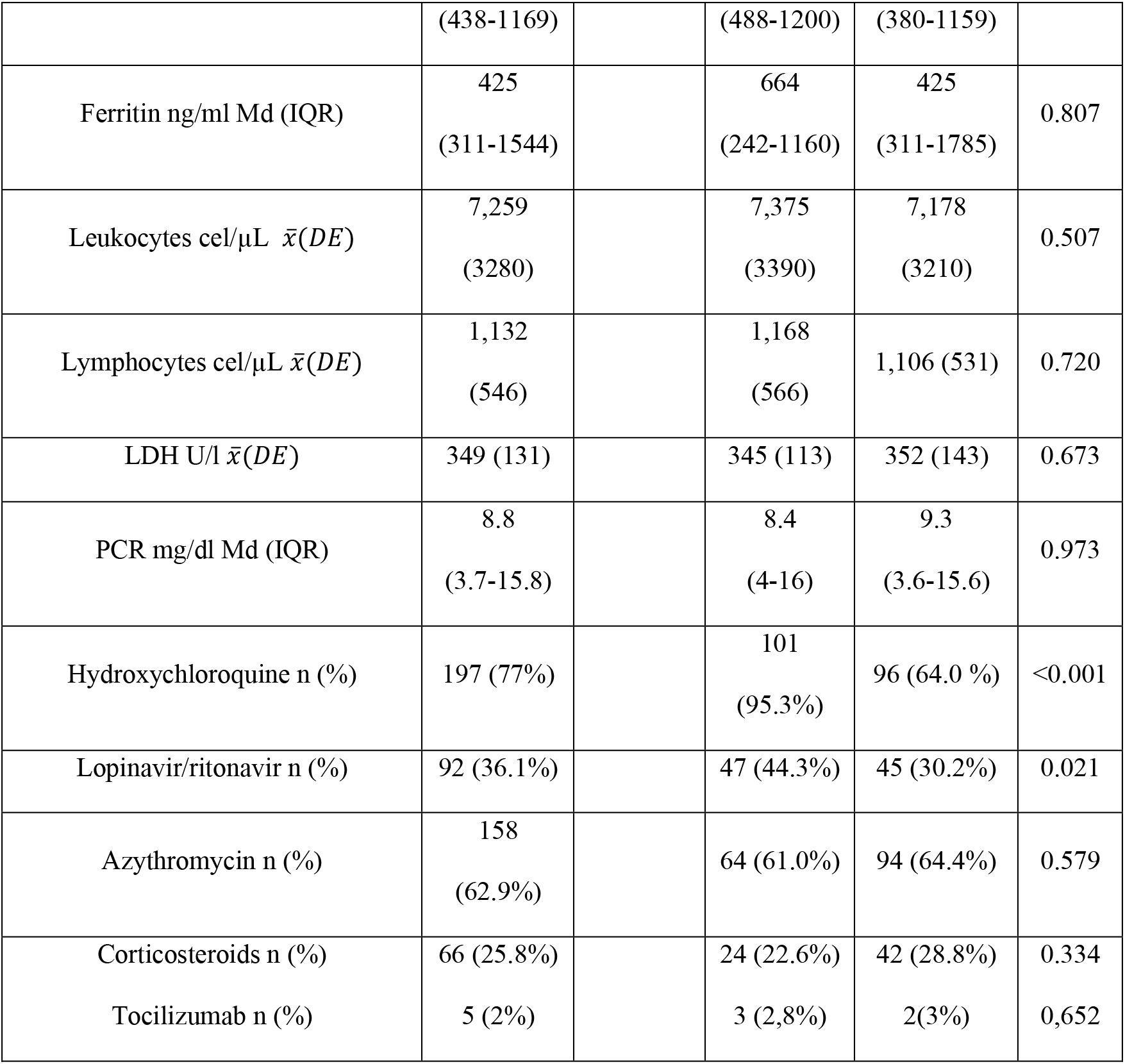
General characteristics of the patients in both treatment groups (with and without INF1β1b).

Of all the patients of our cohort, 106 were treated with interferon beta1b. No significant differences were observed upon comparing the two groups in demographics variables, comorbidities, severity clinical at admission and level of biomarkers at admission. There was more percentage of patients with at least 2 comorbidities in the group not receiving interferon (46.3% vs. 34.9%, p=0.069) (Table 1).

In the interferon group, more patients received hydroxychloroquine (95.3% vs. 64%, p<0.001) and lopinavir/ritonavir (44.3% vs. 30.2%, p=0.021). There was no difference in in-hospital mortality between the group receiving the combination of interferon beta1b and lopinavir/ritonavir versus the group of patients receiving interferon beta1b without lopinavir/ritonavir (p=0.906)

### Outcome

The overall mortality rate was 24.6% (63/256). Twenty-two patients (20.8%) receiving interferon beta1b died and 41 (27.3%) patients not receiving interferon bea1b (p=0.229).

### Mortality risk factors

In the multivariate analysis age (older than 65 years old), clinical severity at admission, and not have received hydroxychloroquine were significantly associated with in-hospital mortality (Table 2). The interferon treatment was not associated with survival benefit neither univariate analysis nor multivariate analysis.

**Table 2.** Multivariate analysis. Risks factors for hospital mortality in COVID-19

|  | **OR** | **95%CI** | **P value** |
| --- | --- | --- | --- |
| Age (<65 years) | 0.002 | 0.00-0.27 | 0.002 |
| Hidroxychloroquine treatment | 0.158 | 0.03-0.79 | 0.025 |
| Clinical severity * |  |  |  |
| Mild | 0.032 | 0.00-0.18 | <0.001 |
| Moderate | 0.089 | 0.01-0.44 | 0.003 |

\* Compared to severe
OR: Odd Ratio; CI: Confidence interval.

## DISCUSSION

Three human coronavirus (SARS-CoV, MERS-CoV and SARS-CoV-2) cause severe pneumonia. Rapid virus replication reaching high titers and associated enhanced inflammatory responses, are believed to contribute to pathogenicity (14 15 16). Type 1 IFNs generally protect mammalian hosts from virus infections, but in some cases, IFN-I is pathogenic. In MERS-CoV–infected mice, IFN-I administration within 1 day after infection protected mice from lethal infection. In contrast, delayed IFN-β treatment failed to inhibit virus replication and enhanced proinflammatory cytokine expression, resulting in fatal pneumonia (17). In that way, it seems that the timing of IFN-I administration is a crucial factor in the effectiveness of the treatment. In an *in vitro* study, the IFN-I pretreatment resulted in a significant reduction in viral replication in Vero E6 cells infected with SARS-CoV-2 in compared to control untreated cells (18).

In our cohort, the median time from the start of symptoms to hospital admission was 7 days in both groups. We do not know if earlier administration would have changed the results of our study. In recently published clinical trial in severe COVID-19, lopinavir/ritonavir treatment within 12 days after the onset of symptoms was associated with shorter time to clinical improvement but later it was not (19).

In our cohort, in-hospital mortality in patients not receiving interferon beta1b was higher than in the interferon group, although the difference was not significant. Most patients in both treatment groups were receiving other potentially effective drugs, as hydroxychloroquine or azytromycin, making it difficult to demonstrate efficacy of a treatment individually in a retrospective analysis. Results from in vitro and animal studies suggest that a combination of lopinavir/ritonavir and interferon-β1b may be effective against MERS-CoV. A randomized controlled trial is ongoing to evaluate this combination as a potential treatment for MERS (MIRACLE trial) (20). To explore the efficacy of this combination in SARS-CoV-2, we compare in-hospital mortality between patients receiving interferon beta1b with and without lopinavir/ritonavir and no difference in in-hospital mortality was found.

There were no significant differences in baseline characteristics between two groups, including the clinical severity of pneumonia on admission. On admission, almost half of the patients presented mild pneumonia in both groups (interferon and control). In fact, 45% of patients who received interferon beta1b during hospitalization presented mild pneumonia on admission. Although treatment with interferon was indicated in moderate-severe pneumonia, there was a shortage of the drug, so not all patients could receive treatment despite being indicated. In this context, interferon beta1b treatment was prioritized based on life expectancy prior to admission. This explains why there were more patients with dementia and other comorbidities in patients not receiving interferon beta1b.

In the multivariate analysis, the predictors of in-hospital mortality were age, severity of pneumonia on admission, and treatment with hydroxychloroquine. The survival benefit found with hydroxychloroquine treatment supports the findings of a previous study of our research group in this sense (under per review). In addition, severity clinical on admission as predictor of mortality is a finding reported in MERS. So, Mohammed Al Ghamdi et al. found in a retrospective cohort study that patients receiving beta interferon and mofetil had improved survival, however this was confounded by the severity of illness on presentation for beta interferon (21).

Our study has the limitations of a retrospective study, selection and unmeasured confounding bias cannot be completely excluded. It would have been ideal to have had microbiological confirmation of all the patients, but since this study had been carried out in the worst phase of the epidemic in our country, at specific times and due to lack of tests, microbiological confirmation was not necessary in cases of high clinical and radiological suspicion without any other alternative cause. Furthermore, we cannot evaluate differences between the two groups in terms of virus elimination, as there was no RT-PCR monitoring for all patients.

This study is, to the best our knowledge, the first clinical study that evaluates the effectiveness of the interferon beta1b in SARS-CoV-2 infection.

### Conclusion

In our retrospective cohort, the interferon beta1b treatment had no significant impact on in-hospital survival. We consider that it would be of interest to explore the efficacy of the early administration of this drug in controlled clinical trials, with a larger sample size.

## Data Availability

NA

## Notes

### Competing Interest Statement

The authors have declared no competing interest.

### Funding Statement

No funded

